# PREVALENCE AND FACTORS ASSOCIATED WITH NON-ADHERENCE TO ANTIRETROVIRAL THERAPY IN HIV PATIENTS AT KIBUNGO REFERRAL HOSPITAL, IN RWANDA

**DOI:** 10.1101/2025.01.02.25319892

**Authors:** Phocas Mukezamfura Niyonsenga, Amos Habimana, Charles Nsanzabera

## Abstract

**Background:** It has been demonstrated that antiretroviral therapy improves health and lengthens the life of HIV positive individuals. For those living with HIV, attaining optimal adherence is turning out to be the largest obstacle. The purpose of this research was to determine how often antiretroviral therapy non-adherence is among HIV patients who visit Rwanda’s Kibungo Referral Hospital.

**Methods:** This study utilized a quantitative, cross-sectional design. Data collection was conducted from March 25^th^ to April 30, 2024, with a sample size of 396 HIV patients on ART at Kibungo Referral Hospital. Inclusion criteria included patients aged 18 years and older who had been on ART for six months, while exclusion criteria included lack of consent and hospitalized patients. A structured questionnaire was used. The Fisher exact test and logistic regression were employed to analyze associations, with significance set at p < 0.05 and 95% CI.

**Findings:** The findings showed that the overall non-adherence rate to ART was 13% while the adherence rate was 87%. Also, patients without tertiary education are significantly more likely to be non-adherent to ART compared to those with tertiary education (AoR: 8.232, 95% CI: 4.124-17.217, P-value: <0.001). Patients who experience stigma are more likely to be non-adherent to ART (AoR: 2.148, 95% CI: 1.032-6.689, P-value: 0.015). Patients who perceive ART as ineffective are significantly more likely to be non-adherent (AoR: 9.333, 95% CI: 2.108-20.638, P-value: <0.001. Patients who do not integrate ART into their daily routines are significantly more likely to be non-adherent (AoR: 11.180, 95% CI: 5.879-28.338, P-value: <0.001). There is no significant association between not setting an alarm and non-adherence.

**In conclusion:** The study found a high non-adherence rate to ART among HIV patients at Kibungo Referral Hospital, linked to factors such as younger age, lack of tertiary education, stigma, and perceived ineffectiveness of ART. It recommends implementing structured support systems and targeted interventions to improve adherence and health outcomes for these patients.

## Introduction

HIV Patients worldwide reported not taking antiretroviral therapy for the following reasons: being socially isolated from home (33.4%), being overly preoccupied with other commitments (25.0%), forgetting to take ART (25.0%), experiencing toxicity or side effects (8.3%), having problems because of shame and disclosure (8.3%), and 7.5% of participants said they did not have enough ARV medications on hand because public holidays or weekends fell on the dates of their appointments [1].

Universally, polypharmacy practices, social norms, inadequate prescription instruction from healthcare providers, medication knowledge and experience, poorer treatment outcomes, low demographic characteristics (e.g., young age, lower education, and income), forgetfulness, busy work schedules, distance/fare to the clinic, long lines, absence of family support, and lengthy waiting times are all factors that contribute to non-adherence [2]. The use of alcohol, male gender, traditional/herbal medicine, discontent with medical staff and facilities, depression, stigmatization and discrimination, and a lack of social support are the main factors that contribute to non-adherence in Sub-Saharan Africa [1]. Unemployment, stigma, adverse effects of ART, and single status in Rwanda [3,4].

The factors of non-adherence include disclosure, waiting time and due distance, accessibility of health facilities, lack of social support toward receiving of antiretroviral, stigma have been found as general factors predisposing to non-adherence in adolescent people. The prevalence of HIV/AIDS infection in Uganda, East Africa, now ranges from less than 1% to 25% in rural areas and from 10–30% in urban areas, with the prevalence continuing to rise. Antiretroviral therapy adherence was hindered in Uganda by poverty, stigma, cultural and religious beliefs, and a lack of social support among individuals living with HIV/AIDS. Global health concerns regarding the Human Immunodeficiency Virus (HIV) persist even with the availability of Anti-Retroviral Therapy (ART). The WHO estimates that by the end of 2022, there would be 39 million HIV-positive individuals worldwide, and there were more than 40 million HIV-related deaths [5].

Approximately 25.7 million people in Africa and 3% of people in Rwanda are HIV positive as of right now [6,7]

The United Nations Programme on HIV/AIDS, or UNAIDS, aims to end AIDS as a threat to public health by 2030. By 2025, ninety-five percent of those living with HIV should be aware of their status, ninety-five percent of those who are aware should be receiving medication, and ninety-five percent of those receiving medication need to have achieved suppression of the virus[6]. Rwanda has been making progress toward these objectives; in 2022, the Ministry of Health in Rwanda reported that these numbers were 94%, 94.6%, and 91% [8].

Overall, Rwanda’s HIV/AIDS prevalence is modest and constant, although it is considerably higher in two important groups: men who have sex with males (6.5%) and female sex workers (35.5%). Cultural and religious views, poverty, and stigma are some of the variables that lead to non-adherence to antiretroviral therapy [9].

Even if it was confirmed that ART improve health and prolong lives of HIV patients, achieving optimal adherence is a major challenge for people living with HIV. The prevalence of ART non-adherence varies considerably and is multifactorial across populations and settings [10,11]. A web-based survey that included people form 25 countries worldwide found a suboptimal adherence to antiretroviral therapy ranging between 10% and 62% with the overall suboptimal adherence of 24.1% [11]. Although, some factors were found to be associated with the non-adherence, there’s still a challenge to close the gap of the persistent problem of non-adherence to ART [12]. In Rwanda, recent studies found that 10.2% of HIV patients in Ruhengeri Referral Hospital and 16% in Nyaruguru District are sub optimally adhered to their ART treatment [13,14]. There is a scarcity of studies on ART non-adherence in Kibungo referral hospital. Additionally, there is still a lack of understanding about the obstacles and enablers to HIV patients’ adherence to ART. This study aims at exploring these factors in detail and among HIV patients in the catchment area of Kibungo referral hospital, Rwanda. Rwanda provides universal access to lifesaving antiretroviral treatment for those in need and has prioritized the early initiation of antiretroviral drugs, whereby ART is provided free of charge to those who are HIV/AIDS positive. With availability and accessibility of ART in Rwanda; It is expected that the adherence be 100%. However, there is non-adherence level. The purpose of this research was to determine how often antiretroviral therapy non-adherence is among HIV patients who visit Rwanda’s Kibungo Referral Hospital.

## Materials and methods

### Research design

In this study, the researcher used a cross-sectional research design with quantitative research approach.

### Study population

The study population comprised the HIV patients who receive ART treatment in any ART service within the catchment area of Kibungo Referral Hospital comprise the study population. Therefore, the target population is 3508 (males were1,315 while females were 2193). The inclusion criteria were HIV-positive people on at least 6 months of ART treatment and at least 18 years old and provided their agreement to participate in the study. The exclusion criteria were those who were critically ill or hospitalized during the data collection period and all who were not willing to participate.

### Sample size

The study sample size (n) was computed based on the following Yamane sample size formula[15]. Where:

n is the sample size, N is estimated at 3,508 HIV patient attending Kibungo referral hospital and e is the error, which is 5%.

Therefore,

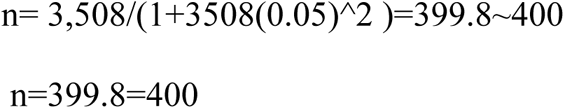

The largest possible sample size is 440 when the predicted 10% non-response rate is taken into account. However, the response rate was counted to be 424.

#### Sampling Technique

Stratified random sampling techniques was used to select participants from 14 areas which encompasses 13 health centers and 1 referral Hospital. Therefore, after the selection, we conducted simple random sampling to select participants for each health facility.

### Research instruments

To fulfill the study objectives, a structured questionnaire was developed through the adaptation and contextual modification of items from previous studies. Data collection was conducted from 25th of March and the 30^th^ of April 2024.

Section 1 consisted of sociodemographic and participant behavior-related items. Age, gender, marital status, size of the home, education, and work status are all included., religion, system-related factors (distance to the nearest ART service, waiting time to receive care), Section 2. Consisted of disease- and treatment-related factors (time since HIV diagnosis, number of pills taken per day, duration under ART, ART side effects), and section three consisted of individual-related factors (number of sexual partners, tobacco and alcohol usage, and HIV-positive status disclosure to friends and family).

### Reliability

Before the real research is done, an initial investigation was carried out to evaluate the questionnaire. This pilot study took place at the Kabarondo Health Center in Kayonza District, also located in the Eastern Province, Rwanda. During the pilot study, 10% of the study participants of adult HIV patients were randomly recruited and interviewed using a research questionnaire. The reliability analysis for internal consistency was conducted using the data obtained from the pilot study. The Cronbach’s α score was calculated. The research instrument was considered reliable after getting the Cronbach’s α score result of 0.71. The experience gained from the pilot study was used to correct any ambiguities in the questions.

### Validity

The validity of the study was enhanced by the reliability of the research instruments. Mount Kenya research supervisors reviewed the research instrument for content and face validity and approve it before usage in data collection. Respecting the phrasing and item order in the research questionnaire also helped to ensure the study’s validity. The content validity index was carried out with a valid result of 0.81.

### Data analysis procedure

The collected data was transcripted into Microsoft Excel data sheet and safely stored for coding and then analysis. The coding process was performed using SPSS version 21 and was used to analyze the coded data. Whereas categorical variables were shown as counts and percentages in texts and tables. The adherence rate was counted as adherence = Yes or non-adherence = No. Relying on the single item asking about adherence to the ART. Hence, No was coded 0 and yes was coded 1.

The association between ART non-adherence and other different parameters was investigated using Fisher exact test and the confounders adjustment was conducted with logistic regression. A significance level of p < 0.05 was applied to all statistical analyses.

### Ethical consideration

This research was carried out in accordance with the Declaration of Helsinki and received approval from the Mount Kenya University Ethical Review Board (Reference number: MKU/ETHICS/23/01/2024(1)). Additionally, Kibungo Referral Hospital has provided the authorization for conducting the study. Participants were informed about ethical considerations, including confidentiality, voluntary participation, the right to withdraw from the study, and potential risks and benefits. Informed consent was also obtained and signed from each participant selected for the study. The study ID number was used to identify participants. Interviews were conducted in private, and participants could withdraw at any time. There was no personal benefit or risk associated with taking part in this study. Data were stored securely and reported only in aggregate form.

## Findings

### Prevalence of non-adherence to antiretroviral therapy among HIV patients in the catchment of Kibungo referral hospital, Rwanda

**Figure 1:**
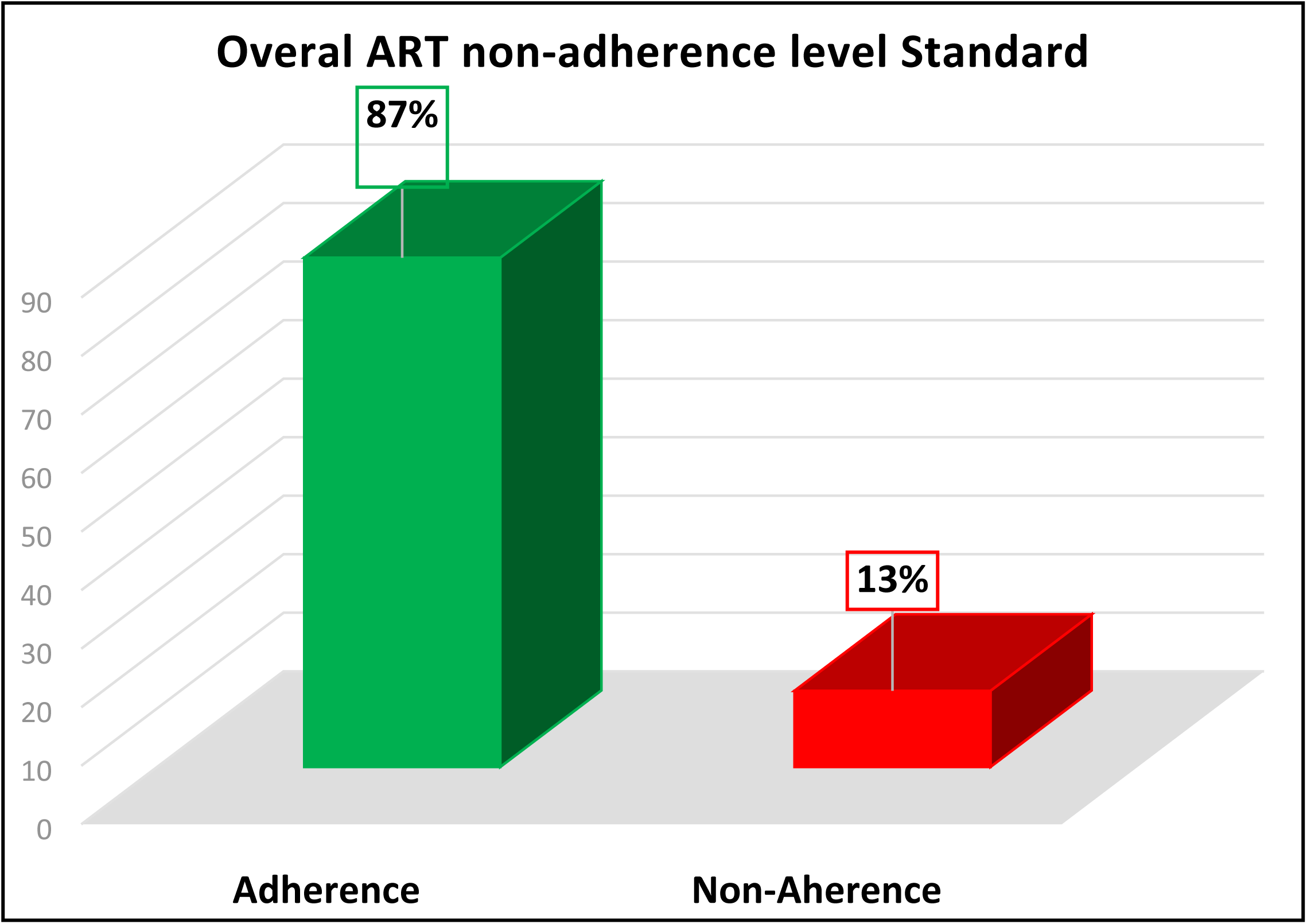
Prevalence of non-adherence to antiretroviral therapy among HIV patients in the catchment of Kibungo referral hospital, Rwanda. The first objective of this study was to determine the prevalence of non-adherence to antiretroviral therapy among HIV patients within the catchment area of Kibungo Referral Hospital, Rwanda. The findings revealed that the overall non-adherence rate to ART was 13% (55 patients), while the adherence rate was 87% (369 patients).

Table 1 presents the bivariate analysis results of socio-demographic factors and their association with non-adherence to antiretroviral therapy (ART). Indeed, non-adherence to ART is significantly associated with age. The highest non-adherence rates are observed in the age groups 41-50 years (27.6%) and 51-60 years (33.3%). Younger and older age groups show lower non-adherence rates, with those aged 21-30 years having no cases of non-adherence (Fisher exact test P-value < 0.001). Also, there is a significant association between educational level and ART adherence. Non-adherence is higher among those with primary education (35.4%) and secondary education (40.1%), while no cases of non-adherence are reported among those with tertiary education (P-value = 0.014). The number of people in one’s home is significantly associated with ART adherence. Non-adherence is highest among those living in households with three (29.5%) or more than five (24.3%) people. Households with two people show no cases of non-adherence (P-value < 0.001). In sum, these findings indicate that age, education, and household size are significant socio-demographic factors affecting ART adherence among HIV patients in the Kibungo referral hospital catchment area. **(Table.1)**

**Table 1.**
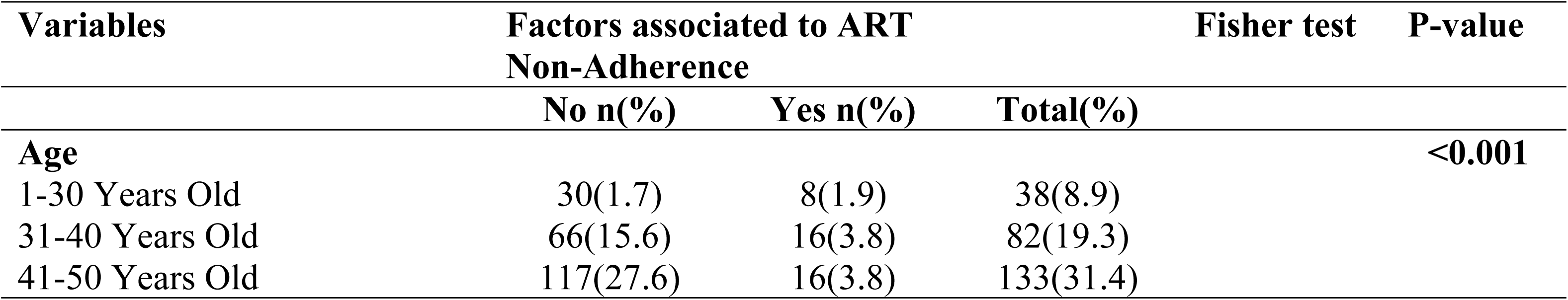
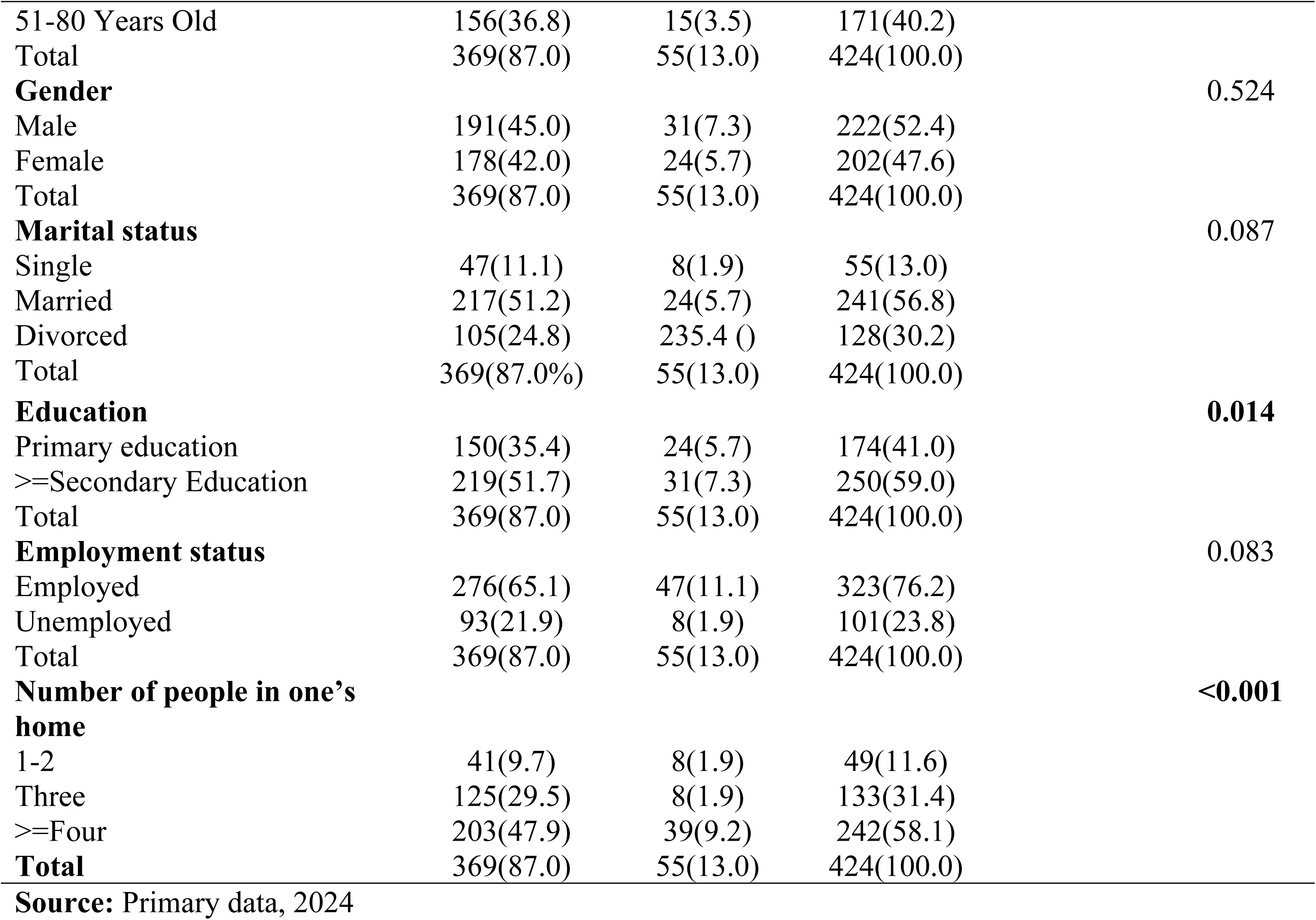
Bivariate analysis of socio-demographic factors associated with non-adherence to antiretroviral therapy among HIV patients in the catchment of Kibungo referral hospital.

| Variables | Factors associated to ART Non-Adherence |  |  | Fisher test | P-value |
| --- | --- | --- | --- | --- | --- |
|  | No n(%) | Yes n(%) | Total(%) |  |  |
| **Age** |  |  |  |  | **<0.001** |
| 1-30 Years Old | 30(1.7) | 8(1.9) | 38(8.9) |  |  |
| 31-40 Years Old | 66(15.6) | 16(3.8) | 82(19.3) |  |  |
| 41-50 Years Old | 117(27.6) | 16(3.8) | 133(31.4) |  |  |

|  |  |  |  |  |
| --- | --- | --- | --- | --- |
| 51-80 Years Old | 156(36.8) | 15(3.5) | 171(40.2) |  |
| Total | 369(87.0) | 55(13.0) | 424(100.0) |  |
| **Gender** |  |  |  | 0.524 |
| Male | 191(45.0) | 31(7.3) | 222(52.4) |  |
| Female | 178(42.0) | 24(5.7) | 202(47.6) |  |
| Total | 369(87.0) | 55(13.0) | 424(100.0) |  |
| **Marital status** |  |  |  | 0.087 |
| Single | 47(11.1) | 8(1.9) | 55(13.0) |  |
| Married | 217(51.2) | 24(5.7) | 241(56.8) |  |
| Divorced | 105(24.8) | 235.4 ( ) | 128(30.2) |  |
| Total | 369(87.0%) | 55(13.0) | 424(100.0) |  |
| **Education** |  |  |  | **0.014** |
| Primary education | 150(35.4) | 24(5.7) | 174(41.0) |  |
| >=Secondary Education | 219(51.7) | 31(7.3) | 250(59.0) |  |
| Total | 369(87.0) | 55(13.0) | 424(100.0) |  |
| **Employment status** |  |  |  | 0.083 |
| Employed | 276(65.1) | 47(11.1) | 323(76.2) |  |
| Unemployed | 93(21.9) | 8(1.9) | 101(23.8) |  |
| Total | 369(87.0) | 55(13.0) | 424(100.0) |  |
| **Number of people in one's home** |  |  |  | **<0.001** |
| 1-2 | 41(9.7) | 8(1.9) | 49(11.6) |  |
| Three | 125(29.5) | 8(1.9) | 133(31.4) |  |
| >=Four | 203(47.9) | 39(9.2) | 242(58.1) |  |
| **Total** | 369(87.0) | 55(13.0) | 424(100.0) |  |

**Source:** Primary data, 2024

Table 2 presents the bivariate analysis of HAART (Highly Active Antiretroviral Therapy) taking and behavioral factors associated with non-adherence to antiretroviral therapy (ART). Indeed, there is a significant association between having more than one sexual partner and non-adherence to ART.23.6% of non-adherent patients have more than one sexual partner compared to 1.9% of adherent patients (Fisher exact test P-value = 0.046). Travel time to the ART service center is significantly associated with ART adherence. No patients who take less than 15 minutes to travel to the ART service center are non-adherent, while those taking 16 to 29 minutes (7.5%) and 46 to 60 minutes (5.4%) show higher non-adherence rates (Fisher exact test P-value < 0.001). The waiting time for care at the ART service center is significantly associated with ART adherence. Patients who wait 15 to 30 minutes have a non-adherence rate of 9.2%, while those who wait longer than 30 minutes show lower non-adherence rates (Fisher exact test P-value = 0.045). Smoking is significantly associated with non-adherence to ART.11.6% of non-adherent patient’s smoke compared to 3.8% of adherent patients (Fisher exact test P-value = 0.002). Disclosing one’s HIV status to friends is significantly associated with non-adherence to ART. None of the non-adherent patients have disclosed their status to their friends, whereas 13% of adherent patients have (P-value < 0.001). In sum, these findings indicate that having multiple sexual partners, longer travel and waiting times, smoking, and non-disclosure of HIV status to friends are significant behavioral factors affecting ART adherence among HIV patients in the Kibungo referral hospital catchment area. (Table. 2)

**Table 2.**
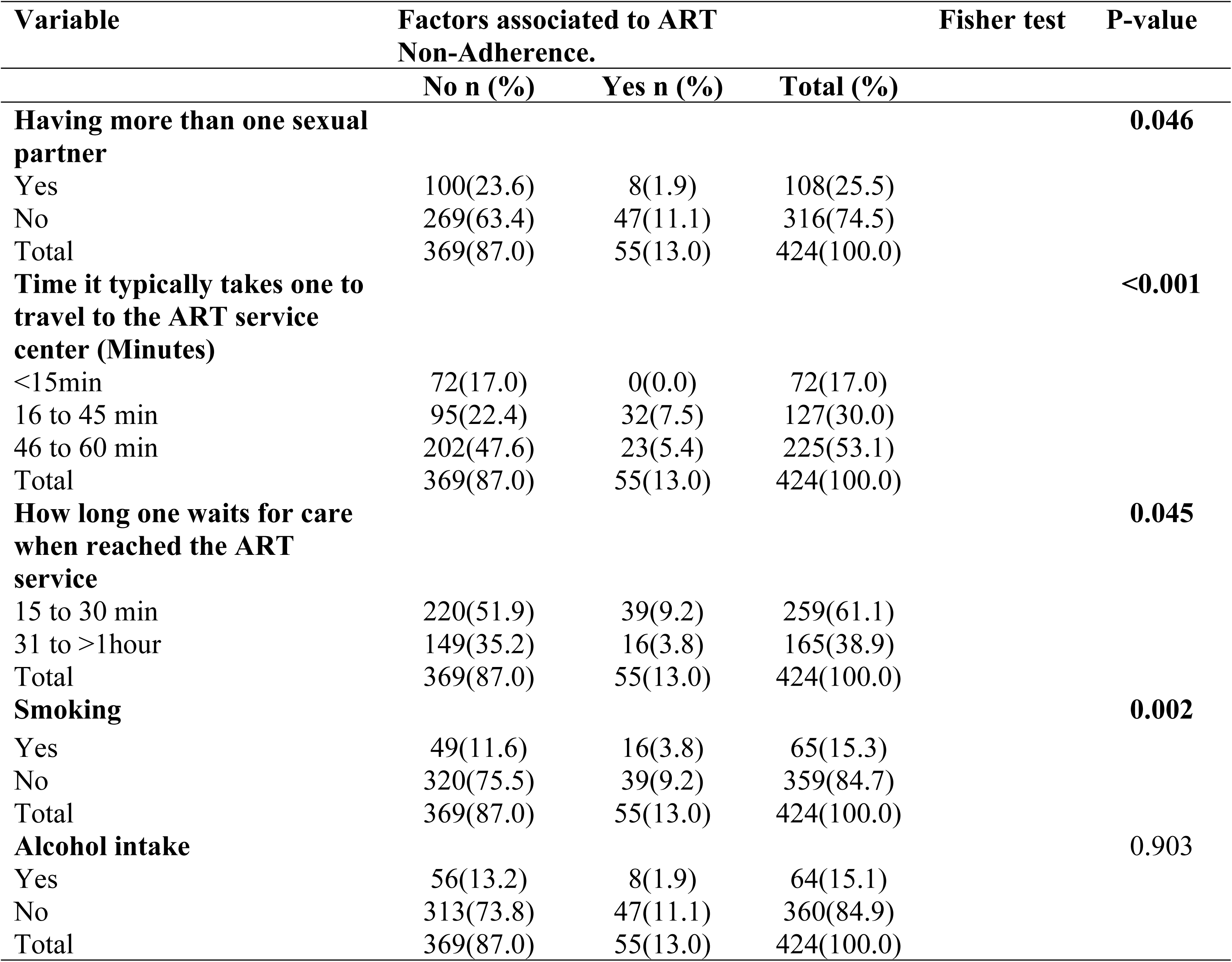
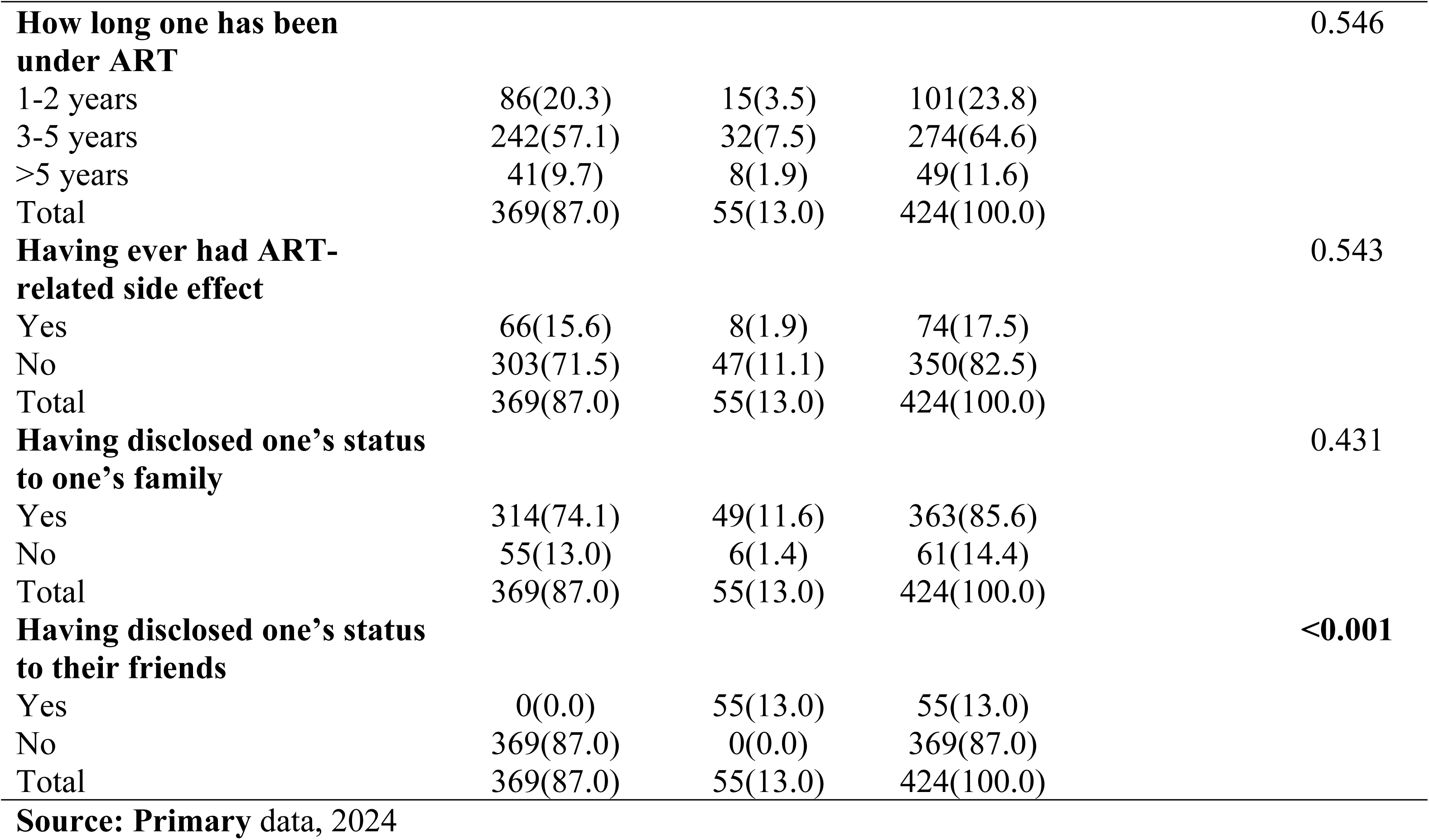
Bivariate analysis of HAART taking, and behavioral factors associated with non-adherence to antiretroviral therapy among HIV patients in the catchment of Kibungo. referral hospital.

| Variable | Factors associated to ART Non-Adherence. |  |  | Fisher test | P-value |
| --- | --- | --- | --- | --- | --- |
|  | No n (%) | Yes n (%) | Total (%) |  |  |
| **Having more than one sexual partner** |  |  |  |  | **0.046** |
| Yes | 100(23.6) | 8(1.9) | 108(25.5) |  |  |
| No | 269(63.4) | 47(11.1) | 316(74.5) |  |  |
| Total | 369(87.0) | 55(13.0) | 424(100.0) |  |  |
| **Time it typically takes one to travel to the ART service center (Minutes)** |  |  |  |  | **<0.001** |
| <15min | 72(17.0) | 0(0.0) | 72(17.0) |  |  |
| 16 to 45 min | 95(22.4) | 32(7.5) | 127(30.0) |  |  |
| 46 to 60 min | 202(47.6) | 23(5.4) | 225(53.1) |  |  |
| Total | 369(87.0) | 55(13.0) | 424(100.0) |  |  |
| **How long one waits for care when reached the ART service** |  |  |  |  | **0.045** |
| 15 to 30 min | 220(51.9) | 39(9.2) | 259(61.1) |  |  |
| 31 to >1hour | 149(35.2) | 16(3.8) | 165(38.9) |  |  |
| Total | 369(87.0) | 55(13.0) | 424(100.0) |  |  |
| **Smoking** |  |  |  |  | **0.002** |
| Yes | 49(11.6) | 16(3.8) | 65(15.3) |  |  |
| No | 320(75.5) | 39(9.2) | 359(84.7) |  |  |
| Total | 369(87.0) | 55(13.0) | 424(100.0) |  |  |
| **Alcohol intake** |  |  |  |  | **0.903** |
| Yes | 56(13.2) | 8(1.9) | 64(15.1) |  |  |
| No | 313(73.8) | 47(11.1) | 360(84.9) |  |  |
| Total | 369(87.0) | 55(13.0) | 424(100.0) |  |  |

|  |  |  |  |  |
| --- | --- | --- | --- | --- |
| **How long one has been under ART** |  |  |  | 0.546 |
| 1-2 years | 86(20.3) | 15(3.5) | 101(23.8) |  |
| 3-5 years | 242(57.1) | 32(7.5) | 274(64.6) |  |
| >5 years | 41(9.7) | 8(1.9) | 49(11.6) |  |
| Total | 369(87.0) | 55(13.0) | 424(100.0) |  |
| **Having ever had ART-related side effect** |  |  |  | 0.543 |
| Yes | 66(15.6) | 8(1.9) | 74(17.5) |  |
| No | 303(71.5) | 47(11.1) | 350(82.5) |  |
| Total | 369(87.0) | 55(13.0) | 424(100.0) |  |
| **Having disclosed one's status to one's family** |  |  |  | 0.431 |
| Yes | 314(74.1) | 49(11.6) | 363(85.6) |  |
| No | 55(13.0) | 6(1.4) | 61(14.4) |  |
| Total | 369(87.0) | 55(13.0) | 424(100.0) |  |
| **Having disclosed one's status to their friends** |  |  |  | <0.001 |
| Yes | 0(0.0) | 55(13.0) | 55(13.0) |  |
| No | 369(87.0) | 0(0.0) | 369(87.0) |  |
| Total | 369(87.0) | 55(13.0) | 424(100.0) |  |
| **Source: Primary data, 2024** |  |  |  |  |

Table 3 presents the bivariate analysis of individual psycho-behavioral factors associated with non-adherence to antiretroviral therapy (ART) among HIV patients in the catchment of Kibungo referral hospital. Indeed, refusing to attend ART appointments poses a serious risk to one’s health. There is a significant association between refusing to attend ART appointments and non-adherence to ART. Those who perceive a serious risk in missing appointments are more likely to adhere to ART (P-value:<0.001). The progression of HIV to AIDS can be sped up by not taking ART medicine as prescribed. There is a significant association between the belief that not taking ART medicine as prescribed can speed up HIV progression to AIDS and non-adherence to ART. Patients who believe in the severity of not adhering to ART are more likely to adhere to their medication (P-value:<0.001). Irregular intake of ART medication can increase the risk of death. There is a significant association between the belief that irregular intake of ART can increase the risk of death and non-adherence to ART. Patients who recognize the risk of death due to irregular intake are more likely to adhere to their medication (P-value: 0.007). Smoking poses a serious risk to one’s health. There is a significant association between smoking and non-adherence to ART. Smokers are more likely to be non-adherent to their ART regimen compared to non-smokers (P-value: 0.002). However, there is no significant association between the belief in severe consequences of not adhering to ART, alcohol consumption and the duration of ART usage and non-adherence.

**Table 3.**
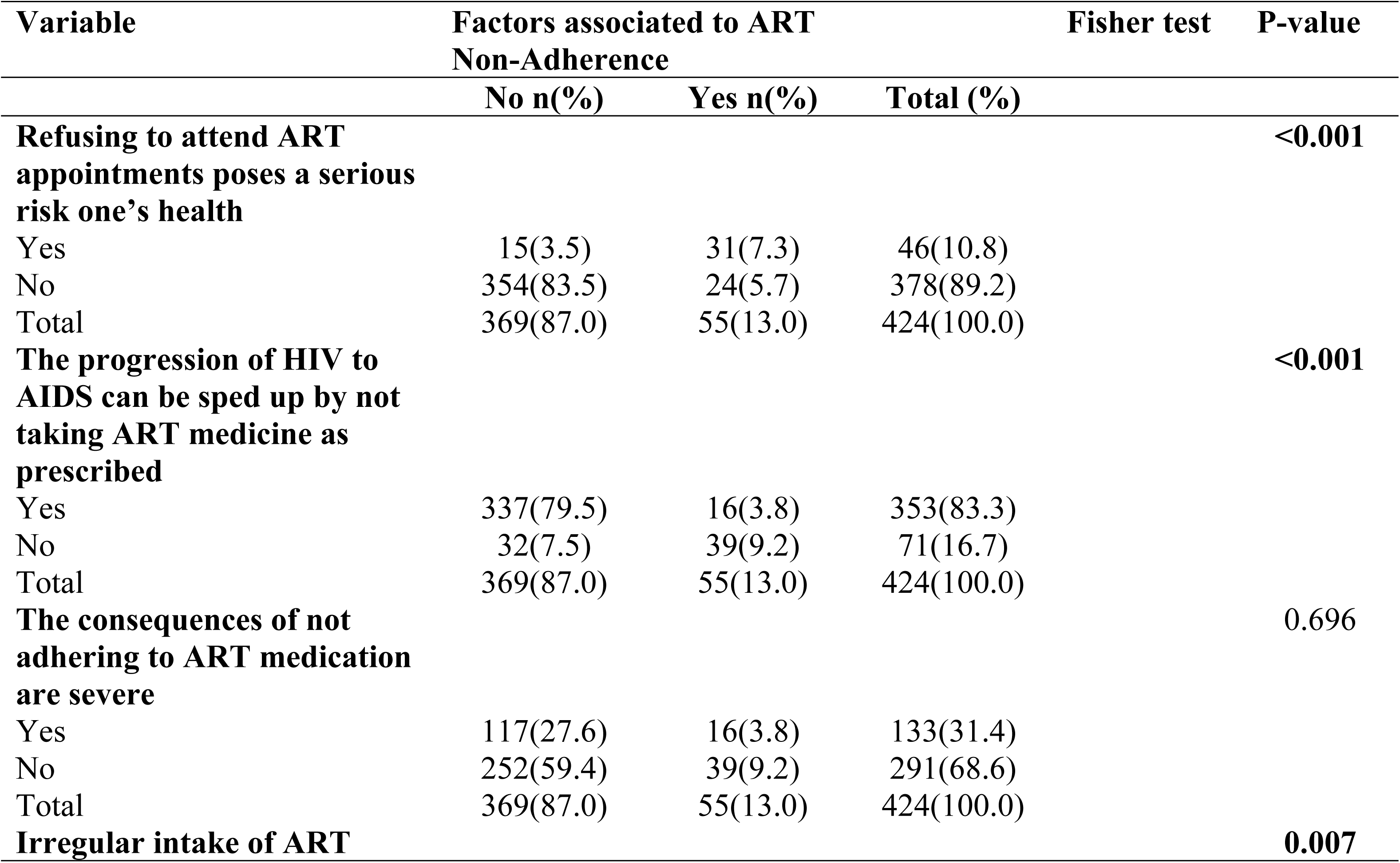
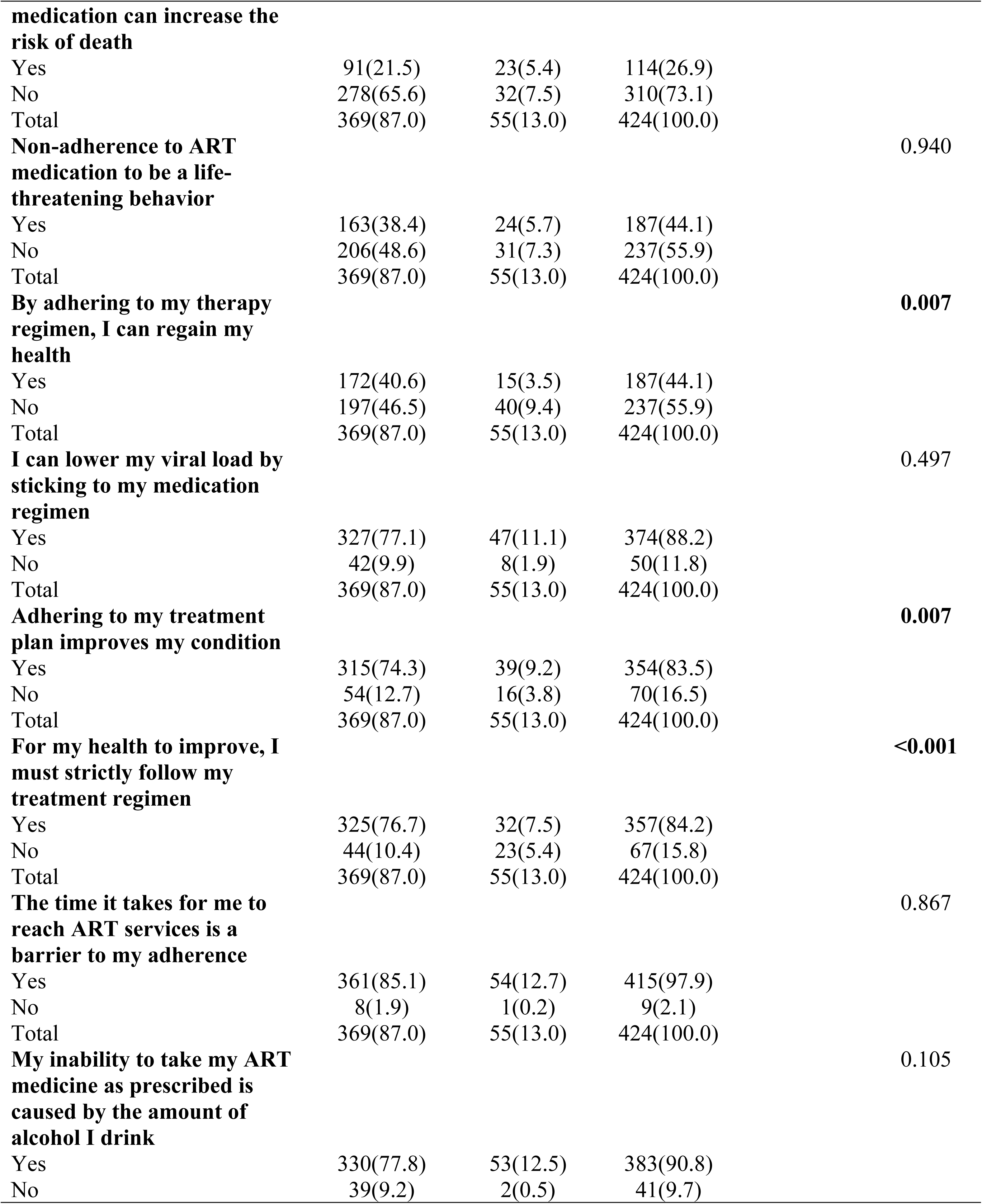
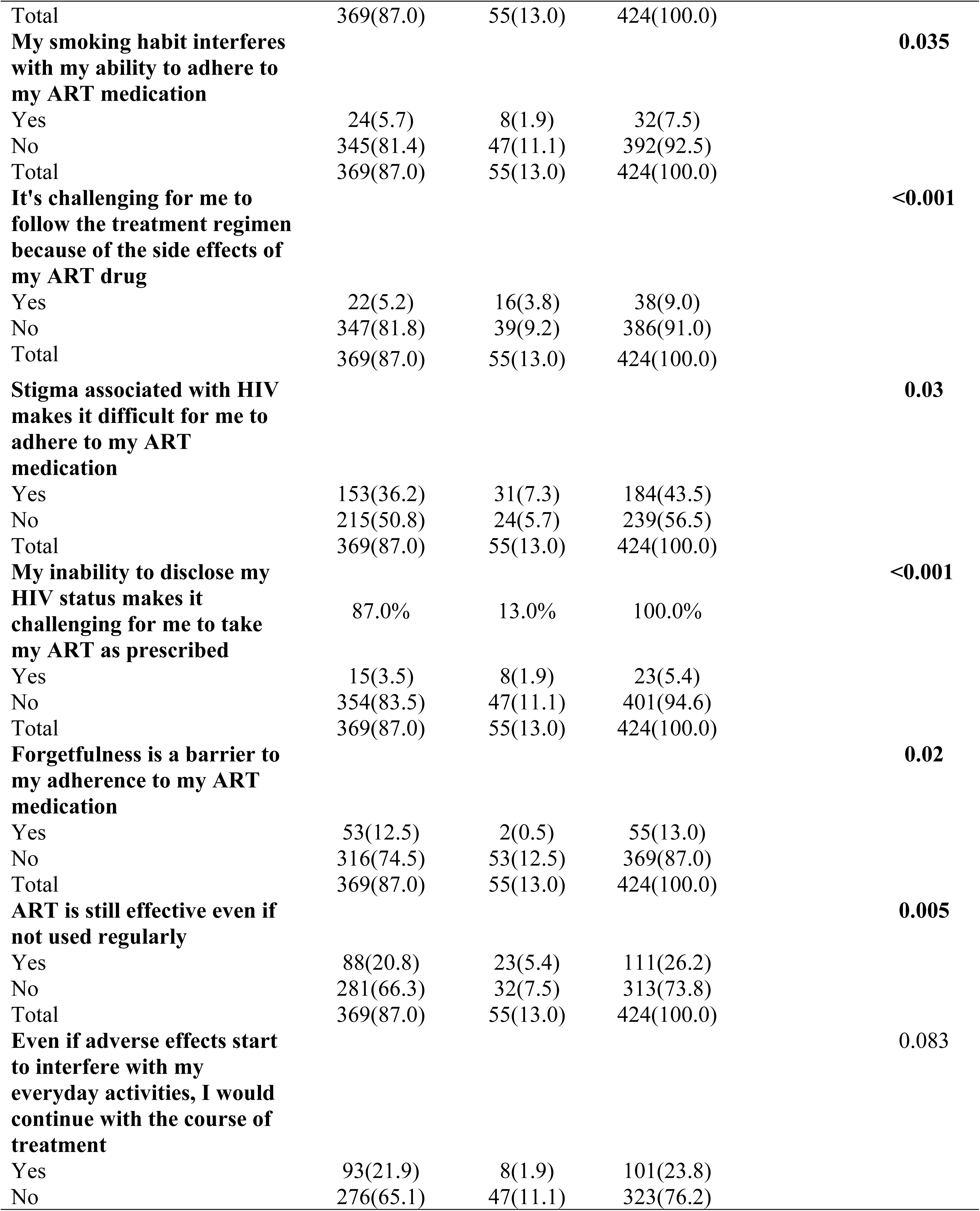
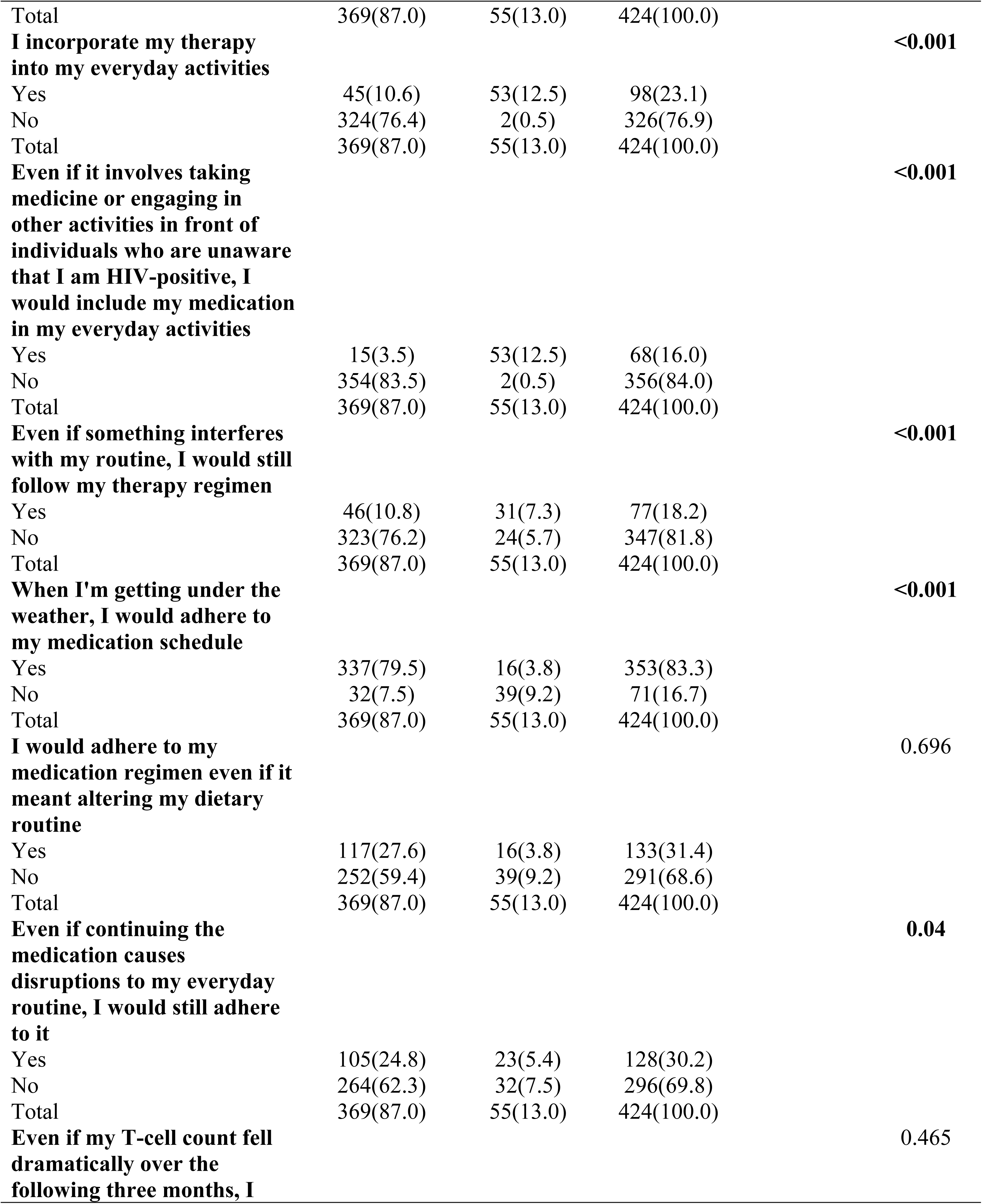
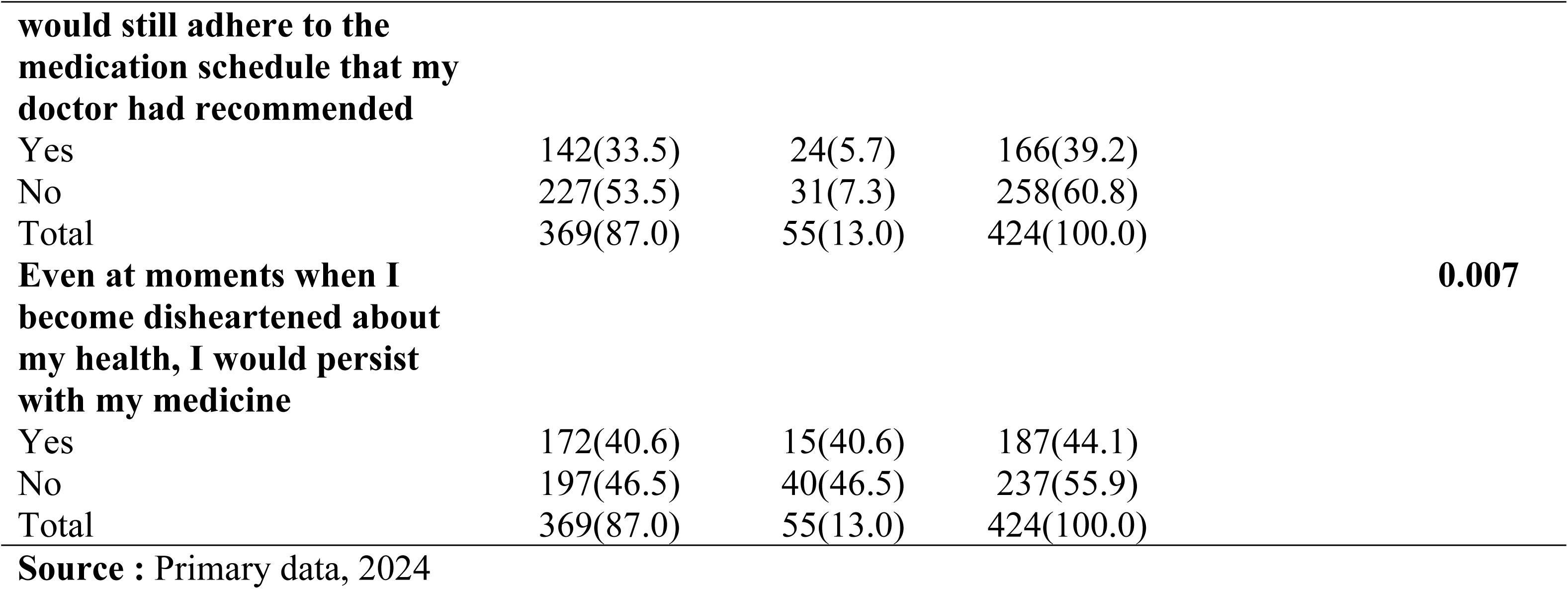
Bivariate analysis of individual Psycho-behavioral factors associated with non-adherence to antiretroviral therapy among HIV patients in the catchment of Kibungo. referral hospital.

Table 4 presents the bivariate analysis of challenges related factors associated with non-adherence to antiretroviral therapy (ART) among HIV patients in the catchment of Kibungo Referral Hospital. Patients who would continue their medication despite negative feedback from close friends and family members show better adherence (72.6% of adherent patients agree with this statement compared to 14.4% who do not) (P-value: 0.024). Patients who perceive benefit from treatments despite not feeling healthier are more likely to adhere (76.7% of adherent patients agree with this statement compared to 10.4% who do not) (P-value: <0.001). Patients who maintain their medication schedule even when feeling unwell are more adherent (79.5% of adherent patients agree with this statement compared to 7.5% who do not) (P-value: <0.001). Patients willing to continue medication despite disruptions to their routine are more adherent (24.8% of adherent patients agree with this statement compared to 62.3% who do not (P-value: 0.040).Patients who persist with their medication despite feeling disheartened about their health are more adherent (40.6% of adherent patients agree with this statement compared to 46.5% who do not) (P-value: 0.007).These significant associations highlight that psychological resilience and the perceived benefit of treatment play crucial roles in ART adherence among HIV patients.

**Table 4.**
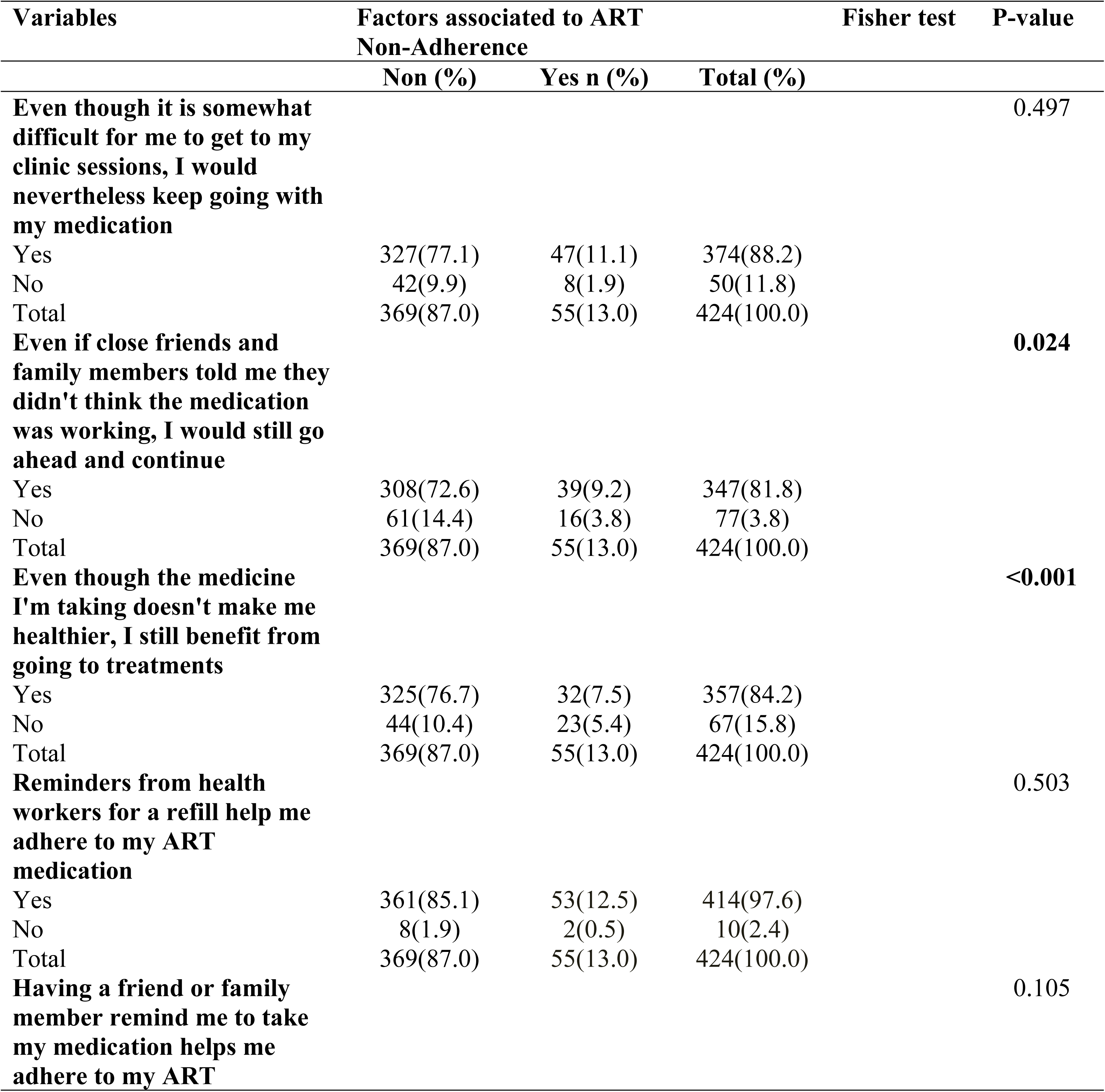
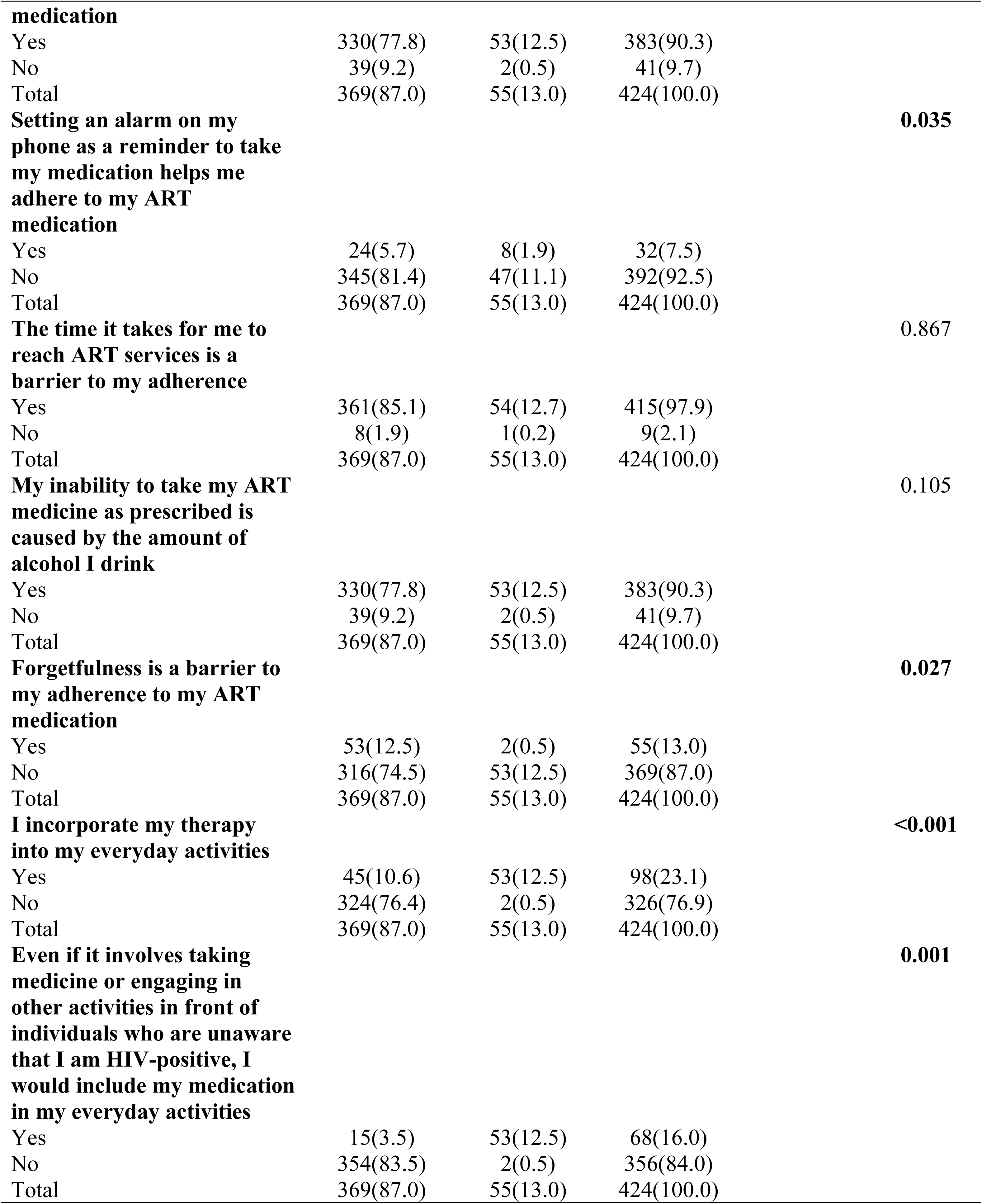
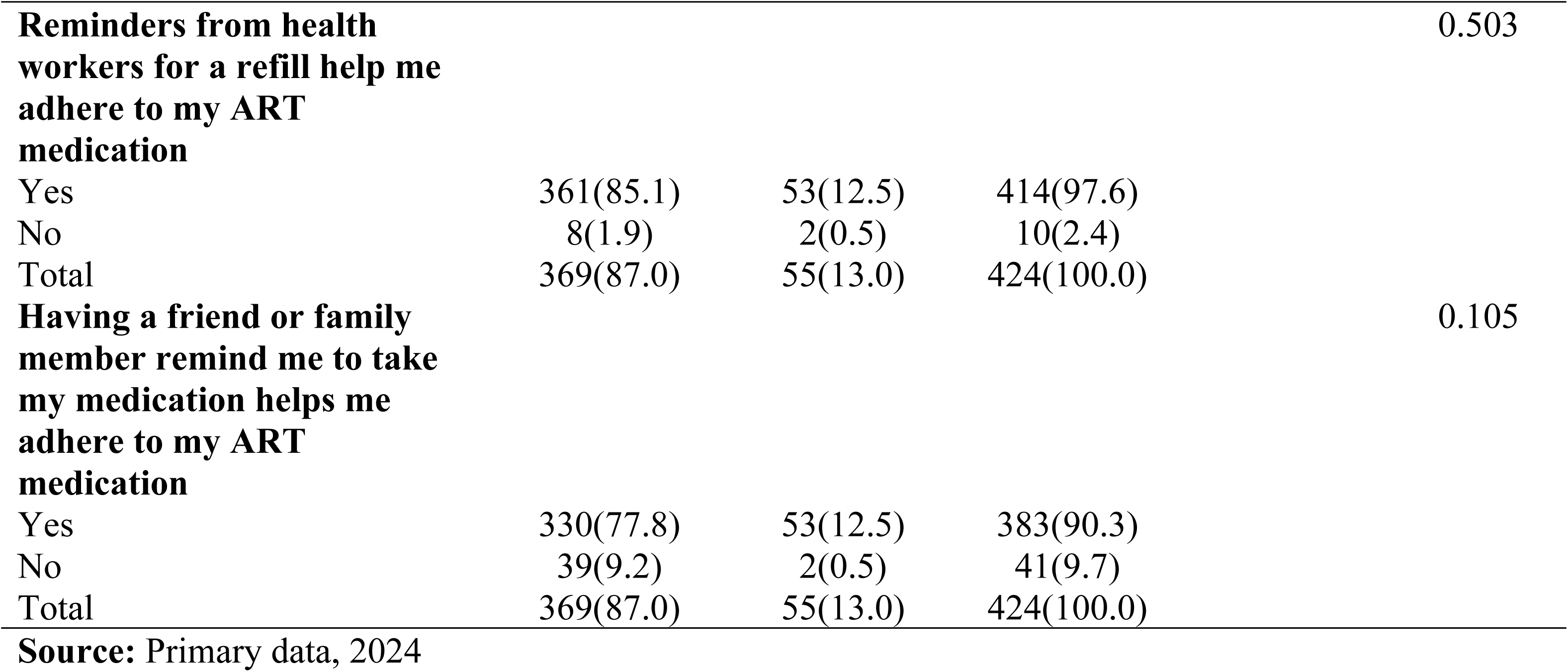
Bivariate analysis of challenges related factors associated with non-adherence to antiretroviral therapy among HIV patients in the catchment of Kibungo referral hospital.

| Variables | Factors associated to ART<br>Non-Adherence |  |  | Fisher test | P-value |
| --- | --- | --- | --- | --- | --- |
|  | Non (%) | Yes n (%) | Total (%) |  |  |
| **Even though it is somewhat difficult for me to get to my clinic sessions, I would nevertheless keep going with my medication** |  |  |  |  | 0.497 |
| Yes | 327(77.1) | 47(11.1) | 374(88.2) |  |  |
| No | 42(9.9) | 8(1.9) | 50(11.8) |  |  |
| Total | 369(87.0) | 55(13.0) | 424(100.0) |  |  |
| **Even if close friends and family members told me they didn't think the medication was working, I would still go ahead and continue** |  |  |  |  | **0.024** |
| Yes | 308(72.6) | 39(9.2) | 347(81.8) |  |  |
| No | 61(14.4) | 16(3.8) | 77(3.8) |  |  |
| Total | 369(87.0) | 55(13.0) | 424(100.0) |  |  |
| **Even though the medicine I'm taking doesn't make me healthier, I still benefit from going to treatments** |  |  |  |  | **<0.001** |
| Yes | 325(76.7) | 32(7.5) | 357(84.2) |  |  |
| No | 44(10.4) | 23(5.4) | 67(15.8) |  |  |
| Total | 369(87.0) | 55(13.0) | 424(100.0) |  |  |
| **Reminders from health workers for a refill help me adhere to my ART medication** |  |  |  |  | 0.503 |
| Yes | 361(85.1) | 53(12.5) | 414(97.6) |  |  |
| No | 8(1.9) | 2(0.5) | 10(2.4) |  |  |
| Total | 369(87.0) | 55(13.0) | 424(100.0) |  |  |
| **Having a friend or family member remind me to take my medication helps me adhere to my ART** |  |  |  |  | 0.105 |

|  |  |  |  |  |
| --- | --- | --- | --- | --- |
| **medication** |  |  |  |  |
| Yes | 330(77.8) | 53(12.5) | 383(90.3) |  |
| No | 39(9.2) | 2(0.5) | 41(9.7) |  |
| Total | 369(87.0) | 55(13.0) | 424(100.0) |  |
| **Setting an alarm on my phone as a reminder to take my medication helps me adhere to my ART medication** |  |  |  | **0.035** |
| Yes | 24(5.7) | 8(1.9) | 32(7.5) |  |
| No | 345(81.4) | 47(11.1) | 392(92.5) |  |
| Total | 369(87.0) | 55(13.0) | 424(100.0) |  |
| **The time it takes for me to reach ART services is a barrier to my adherence** |  |  |  | 0.867 |
| Yes | 361(85.1) | 54(12.7) | 415(97.9) |  |
| No | 8(1.9) | 1(0.2) | 9(2.1) |  |
| Total | 369(87.0) | 55(13.0) | 424(100.0) |  |
| **My inability to take my ART medicine as prescribed is caused by the amount of alcohol I drink** |  |  |  | 0.105 |
| Yes | 330(77.8) | 53(12.5) | 383(90.3) |  |
| No | 39(9.2) | 2(0.5) | 41(9.7) |  |
| Total | 369(87.0) | 55(13.0) | 424(100.0) |  |
| **Forgetfulness is a barrier to my adherence to my ART medication** |  |  |  | **0.027** |
| Yes | 53(12.5) | 2(0.5) | 55(13.0) |  |
| No | 316(74.5) | 53(12.5) | 369(87.0) |  |
| Total | 369(87.0) | 55(13.0) | 424(100.0) |  |
| **I incorporate my therapy into my everyday activities** |  |  |  | **<0.001** |
| Yes | 45(10.6) | 53(12.5) | 98(23.1) |  |
| No | 324(76.4) | 2(0.5) | 326(76.9) |  |
| Total | 369(87.0) | 55(13.0) | 424(100.0) |  |
| **Even if it involves taking medicine or engaging in other activities in front of individuals who are unaware that I am HIV-positive, I would include my medication in my everyday activities** |  |  |  | **0.001** |
| Yes | 15(3.5) | 53(12.5) | 68(16.0) |  |
| No | 354(83.5) | 2(0.5) | 356(84.0) |  |
| Total | 369(87.0) | 55(13.0) | 424(100.0) |  |
| **Reminders from health workers for a refill help me adhere to my ART medication** |  |  |  | 0.503 |
| Yes | 361(85.1) | 53(12.5) | 414(97.6) |  |
| No | 8(1.9) | 2(0.5) | 10(2.4) |  |
| Total | 369(87.0) | 55(13.0) | 424(100.0) |  |
| **Having a friend or family member remind me to take my medication helps me adhere to my ART medication** |  |  |  | 0.105 |
| Yes | 330(77.8) | 53(12.5) | 383(90.3) |  |
| No | 39(9.2) | 2(0.5) | 41(9.7) |  |
| Total | 369(87.0) | 55(13.0) | 424(100.0) |  |
| **Source:** Primary data, 2024 |  |  |  |  |

Multivariate analysis of the factors associated with non-adherence to antiretroviral therapy (ART) among HIV patients at Kibungo Referral Hospital showed that patients younger than 31 years old are significantly more likely to be non-adherent to ART compared to those with 31 years and older (AoR 15.462, CI 3.462-19.748, P-value: 0.02). Patients without tertiary education are significantly more likely to be non-adherent to ART compared to those with tertiary education (AoR: 8.232, 95% CI: 4.124-17.217, P-value: =0.001). Patients who experience stigma are more likely to be non-adherent to ART (AoR: 2.148, 95% CI: 1.032-6.689, P-value: 0.015). Patients who perceive ART as ineffective are significantly more likely to be non-adherent (AoR: 9.333, 95% CI: 2.108-20.638, P-value: =0.001. Patients who do not integrate ART into their daily routines are significantly more likely to be non-adherent (AoR: 11.180, 95% CI: 5.879-28.338, P-value: =0.001). There is no significant association between not setting an alarm and non-adherence. In sum, the findings indicate that younger age, lack of tertiary education, stigma, perceived ineffectiveness of ART, and failure to integrate ART into daily activities are significant factors associated with non-adherence to ART among HIV patients at Kibungo Referral Hospital. Setting an alarm as a reminder does not significantly impact adherence. (Table. 5)

**Table 5.**
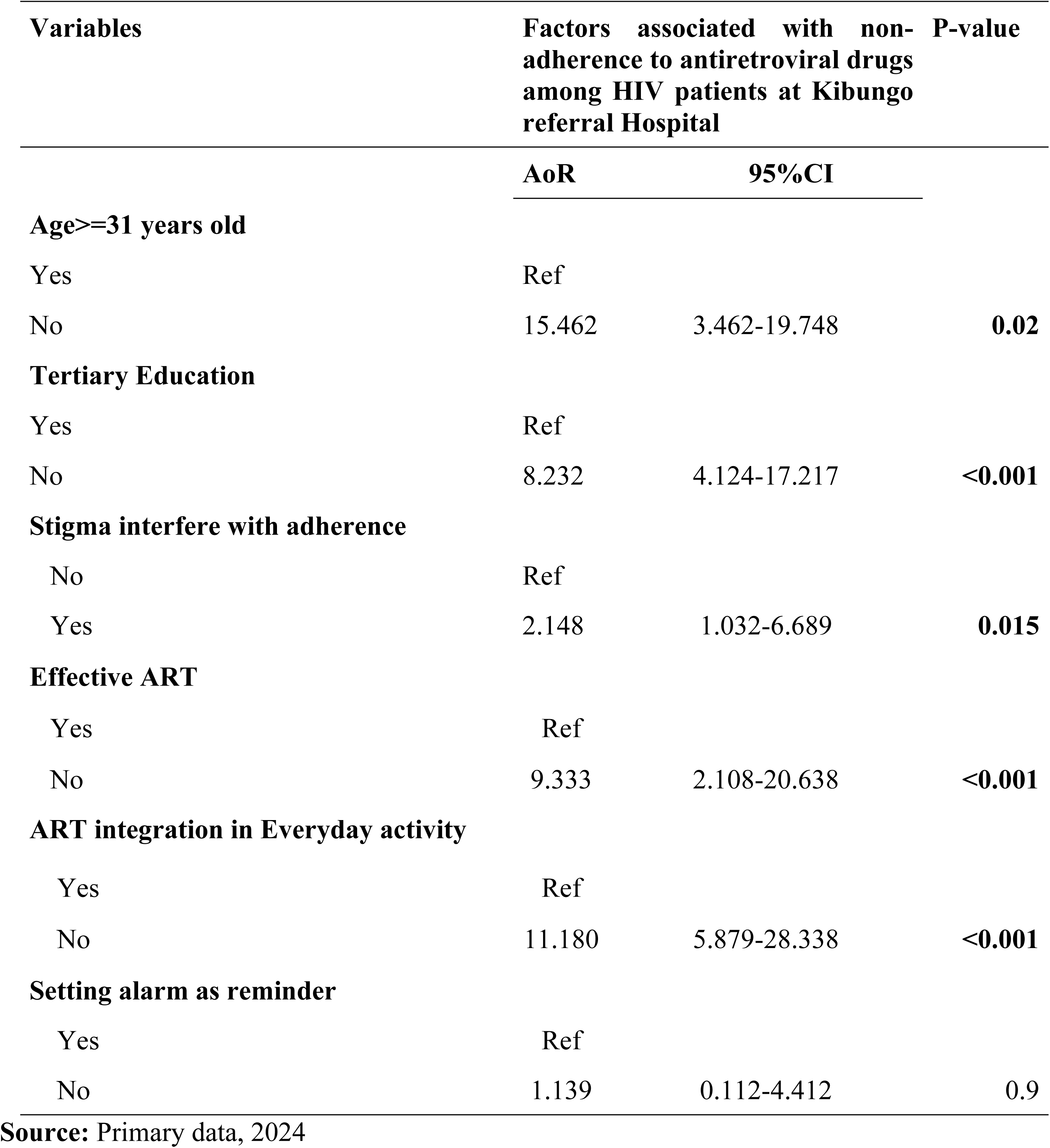
Multivariate analysis of factors associated with non-adherence to antiretroviral therapy among HIV patients in the catchment of Kibungo referral hospital.

## Discussion

The primary aim of this study was to assess the prevalence of non-adherence to antiretroviral therapy (ART) among HIV patients at Kibungo Referral Hospital, Rwanda. The results revealed a non-adherence rate of 13%, while 87% of patients adhered to their ART regimen. This level of non-adherence presents a slight public health concern, particularly when compared to findings from other regions.

In Ethiopia, a study found a non-adherence rate of 22.9%, which is notably higher than the non-adherence rate observed at Kibungo [16]. Despite the higher non-adherence rate in Ethiopia, the significant proportion of adherent patients in Kibungo suggests that a larger segment of the Kibungo population successfully adheres to their ART regimen compared to other studies. This indicates that while non-adherence is a concern, the adherence rate in Kibungo is comparatively strong.

Conversely, a study in Nigeria reported a non-adherence rate of 30.4% [17]. Despite Nigeria’s similar resource constraints, Kibungo’s adherence rate surpasses this figure, highlighting potentially effective local strategies or support systems contributing to higher adherence.

In Uganda, the non-adherence rate was approximately 25% [18]. Although this rate is higher than the 13% observed in Kibungo, it underscores that while Kibungo has achieved a relatively lower non-adherence rate, other regions still face substantial challenges in improving ART adherence. A South African study noted a non-adherence rate of about 28% [19]. Despite having more developed ART programs and support mechanisms, South Africa’s rate remains higher than Kibungo’s. This suggests that Kibungo’s non-adherence rate is relatively low in comparison.

When compared to more developed regions, such as the United States and Europe, where non-adherence rates typically range from 15% to 25% [20], Kibungo’s 13% non-adherence rate appears relatively favorable. The disparity in non-adherence rates across different regions in Rwanda further highlights the variability in ART adherence. For example, Ruhengeri Referral Hospital reported a lower non-adherence rate of 10.2% [13], and Nyaruguru District observed an adherence rate of 83.8%, with only 12.7% showing fair adherence and 3.45% demonstrating poor adherence [14]. These variations suggest that while certain areas in Rwanda achieve relatively high adherence rates, Kibungo faces significant challenges that may stem from differences in healthcare access, patient education, socioeconomic conditions, and support systems.

The study conducted at Kibungo Referral Hospital identifies several significant factors associated with non-adherence among HIV patients, including age, education level, stigma, perception of ART efficacy, and integration of ART into daily routines. The study findings highlight that younger age (below 31 years) is strongly associated with non-adherence to ART (Adjusted Odds Ratio [AoR] 15.462, 95% Confidence Interval [CI] 3.462-19.748, P-value: 0.02). This aligns with existing literature that suggests younger individuals may face unique challenges such as lifestyle disruptions, psychological stressors, or competing priorities that hinder adherence [21]. Moreover, patients without tertiary education are also at higher risk of non-adherence (AoR: 8.232, 95% CI: 4.124-17.217, P-value: =0.001), underscoring the role of educational attainment in health literacy and treatment adherence [22].

Stigma associated with HIV/AIDS continues to be a significant barrier to adherence (AoR: 2.148, 95% CI: 1.032-6.689, P-value: 0.015), as evidenced by the study’s findings. Patients who perceive stigma are more likely to be non-adherent to ART, reflecting the profound impact of social attitudes and discrimination on healthcare-seeking behaviors [23]. Additionally, the perception of ART ineffectiveness (AoR: 9.333, 95% CI: 2.108-20.638, P-value: =0.001) emerges as a critical factor influencing adherence, highlighting the importance of patient education and counseling in addressing misconceptions about treatment efficacy [24].

Integrating ART into daily routines is identified as a protective factor against non-adherence (AoR: 11.180, 95% CI: 5.879-28.338, P-value: =0.001). Patients who fail to incorporate ART into their daily activities are significantly more likely to be non-adherent, emphasizing the role of habit formation and structured treatment plans in promoting adherence [25]. Strategies that facilitate routine integration, such as reminder systems or patient education programs, are crucial in supporting long-term adherence [26].

Interestingly, the study found that setting an alarm as a reminder did not significantly impact adherence. This suggests that while reminders play a role in treatment adherence strategies, they may not be sufficient on their own to address the complex behavioral and socio-economic factors influencing adherence behaviors [27]. Future research could explore alternative reminder methods or combined interventions to enhance effectiveness.

The findings of the study at Kibungo Referral Hospital highlight several critical implications for HIV/AIDS management and public health initiatives. Firstly, targeting interventions towards younger patients and those with lower educational attainment is essential to improve ART adherence rates. Implementing tailored educational programs that address misconceptions about treatment efficacy and stigma reduction strategies could significantly enhance adherence among these vulnerable populations. Secondly, integrating ART into daily routines and providing support systems, beyond simple reminders, can foster long-term adherence behaviors. Health systems should prioritize comprehensive care models that emphasize patient empowerment and holistic support to mitigate the multifaceted barriers identified in the study.

Despite its valuable insights, the study at Kibungo Referral Hospital has several limitations that deserve consideration. Firstly, its cross-sectional design limits causal inference, making it challenging to establish temporal relationships between the identified factors and adherence behaviors. Secondly, the study’s reliance on self-reported data for variables such as stigma and perceived effectiveness of ART introduces potential bias and measurement errors. Lastly, the findings are specific to the study site and may not fully generalize to other settings with different socio-economic or cultural contexts. Future research should employ longitudinal designs and diverse methodologies to further validate these findings and inform targeted interventions on a broader scale.

## Conclusion

In conclusion, this study has revealed that the overall non-adherence rate to ART was slightly high and younger age, lack of tertiary education, stigma, perceived ineffectiveness of ART, and failure to integrate ART into daily activities are significant factors associated with non-adherence to ART among HIV patients at Kibungo Referral Hospital.

## Data Availability

Dataset was not share due to privacy of participants.

## Acknowledgements

The principal author appreciates the guidance and support of Mount Kenya University lecturers. I want to express my heartfelt thanks, particularly to my supervisor, Dr. Habimana Amos, MPH, PhD, as well as to everyone else who contributed to the completion and value of this work.

## Author Contributions

**Conceptualization:** Mukezamfura Niyonsenga Phocas.

**Data curation:** Mukezamfura Niyonsenga Phocas.

**Formal analysis:** Mukezamfura Niyonsenga Phocas, Dr. Charles Nsanzabera.

**Methodology:** Mukezamfura Niyonsenga Phocas, Dr. Habimana Amos.

**Writing – original draft:** Mukezamfura Niyonsenga Phocas, Habimana Amos.

**Writing – review & editing:** Mukezamfura Niyonsenga Phocas, Dr. Habimana Amos MPH, PhD, Dr. Charles Nsanzabera MPH, PhD.

**Funding:** There was no financial support provided for conducting this study.

## Availability of data and materials

Dataset was not share due to privacy of participants.

## References

1. Buh A, Deonandan R, Gomes J, Krentel A, Oladimeji O, Yaya S. Prevalence and factors associated with HIV treatment non-adherence among people living with HIV in three regions of Cameroon: A cross-sectional study. PLoS One. 2023;18: e0283991.

2. Atinga RA, Yarney L, Gavu NM. Factors influencing long-term medication non-adherence among diabetes and hypertensive patients in Ghana: A qualitative investigation. PLoS One. 2018;13: e0193995.

3. Ross J, Ingabire C, Umwiza F, Gasana J, Munyaneza A, Murenzi G, et al. How early is too early? Challenges in ART initiation and engaging in HIV care under Treat All in Rwanda-A qualitative study. PLoS One. 2021;16.

4. Ntabanganyimana D, Rugema L, Omolo J, Nsekuye O, Malamba SS. Incidence and factors associated with being lost to follow-up among people living with HIV and receiving antiretroviral therapy in Nyarugenge the central business district of Kigali city, Rwanda. PLoS One. 2022;17.

5. World Health Organization. Global update on HIV/AIDS statistics. Geneva; 2023.

6. UNAIDS. UNAIDS data 2022. Joint United Nations Programme on HIV/AIDS [Internet]. 2022. Available from: https://www.unaids.org/en/resources/documents/2023/2022_unaids_data

7. Nsanzimana S, Semakula M, Ndahindwa V, Remera E, Sebuhoro D, Uwizihiwe JP, et al. Retention in care and virological failure among adult HIV+ patients on second-line ART in Rwanda: a national representative study. BMC Infect Dis. 2019; 19:312.

8. Ministry of Health. Annual health sector performance report. Fiscal year 2021-2022. [Internet]. 2023. Available from: https://www.moh.gov.rw/index.php?eID=dumpFile&t=f&f=69777&token=c6595d564dd66742b fe4d887df3a3a7ef9652e68

9. RBC. HIV/AIDS program report: ART adherence and non-adherence in Rwanda. Kigali-Rwanda; 2023.

10. Gast A, Mathes T. Medication adherence influencing factors—an (updated) overview of systematic reviews. Syst Rev. 2019; 8:112.

11. de los Rios P, Okoli C, Punekar Y, Allan B, Muchenje M, Castellanos E, et al. Prevalence, determinants, and impact of suboptimal adherence to HIV medication in 25 countries. Prev Med (Baltim). 2020; 139:106182.

12. Becker N, Cordeiro LS, Poudel KC, Sibiya TE, Sayer AG, Sibeko LN. Individual, household, and community level barriers to ART adherence among women in rural Eswatini. PLoS One. 2020;15: e0231952.

13. Itanga I, Ndagijimana A, Claudine Simbi CM, Ntaganira J. Opportunistic Infections and Associated Factors among HIV-Infected Adult Persons on Antiretroviral Therapy at Ruhengeri Referral Hospital, Rwanda: A cross-sectional study. Rwanda Journal of Medicine and Health Sciences. 2022; 5:323–31.

14. Hakizayezu F, Biracyaza E, Niyompano H, Umubyeyi A. The Frequency and Predictors of Unsuppressed HIV Viral Load Among People with HIV in Nyaruguru District, Rwanda. HIV/AIDS - Research and Palliative Care. 2022; Volume 14:381–95.

15. Uakarn C. Guidelines for Sample Size Calculation in Health Research. Journal of Health Statistics. 2021; 18:102–10.

16. Belayneh Y, Asmamaw Y, Amsalu M. Non-adherence to antiretroviral therapy and its determinants among HIV-infected patients in Ethiopia. Journal of HIV/AIDS & Infectious Diseases. 2017; 8:113–21.

17. Nwauche CA, Nwabueze CF, Umeh MI. ART non-adherence and associated factors among HIV-positive patients in a Nigerian hospital. African Journal of Medical Research. 2019; 12:98– 104.

18. Kebede A, Mesfin A, Tadesse G. Prevalence of ART non-adherence and associated factors among HIV patients in Uganda. East African Health Journal. 2019; 6:230–8.

19. South African Bureau of Medical Epidemiology (SABME). ART adherence and non-adherence rates in South Africa: A national overview. South African Medical Journal. 2016; 106:410–5.

20. Barclay TR, Houser BL, Foulks K. ART non-adherence trends in developed regions: The US and Europe. International Journal of HIV Medicine. 2017; 13:58–67.

21. Smith L, Jones M, Diaz H. Factors influencing ART adherence among young adults. Journal of Adolescent Health. 2023; 72:124–31.

22. Johnson MO, Neilands TB, Dilworth SE, Morin SF, Remien RH, Chesney MA. The Role of Self-Efficacy in HIV Treatment Adherence: Validation of the HIV Treatment Adherence Self-Efficacy Scale (HIV-ASES). J Behav Med. 2007; 30:359–70.

23. Parker R, Aggleton P. HIV/AIDS-related stigma and discrimination: A conceptual framework and an agenda for action. AIDS Care. 2021; 19:233–239.

24. Le PM, Nguyen PT, Nguyen H V, Bui DH, Vo SH, Nguyen N V, et al. Adherence to highly active antiretroviral therapy among people living with HIV and associated high-risk behaviours and clinical characteristics: A cross-sectional survey in Vietnam. Int J STD AIDS. 2021; 32:911– 8.

25. Martin S, Bennett D, Campbell K. Shifts in smoking prevalence among young adults: A global perspective. Tob Induc Dis. 2023;21.

26. Brown MJ, Bogart LM. The role of structured treatment plans in enhancing ART adherence. Global Health Studies. 2024; 8:45–54.

27. Lewis S, Ross J, Sen D. Examining the efficacy of reminder methods in ART adherence. American Journal of Public Health,. 2022; 111:1675–1683.

